# SIGLEC5: An immune checkpoint ligand in sepsis

**DOI:** 10.1101/2020.05.30.20117473

**Authors:** Roberto Lozano-Rodríguez, José Avendaño-Ortíz, Karla Montalbán-Hernández, Juan Carlos Ruiz-Rodríguez, Ricardo Ferrer, Alejandro Martín-Quirós, Charbel Maroun-Eid, Juan José González-López, Anna Fàbrega, Verónica Terrón, Carlos del Fresno, Víctor Toledano, Elvira Marín, María Guitiérrez-Fernández, Elisa Alonso-López, Carolina Cubillos-Zapata, Pablo Stringa, Rebeca Pérez de Diego, Pablo Pelegrin, Carlos García-Palenciano, Jaime Valentín, Paloma Gómez-Campelo, Luis A. Aguirre, Eduardo López-Collazo

## Abstract

Sepsis is a global health priority. Despite thorough studies in mice models, its molecular and cellular basis remain unclear and there is no pharmacological effective treatment other than antimicrobial and supportive therapy. During sepsis, T cells exhaustion compromises patients’ outcome, and immune checkpoints (ICs) become crucial players in disease management. Here, a total of 425 patients with systemic inflammatory response criteria and 127 controls were studied. Soluble SIGLEC5 (sSIGLEC5) levels in plasma were higher in patients with sepsis compared to the other groups and even higher in those patients with septic-shock. sSIGLEC5 plasma levels were higher in non-survivors than in survivors and ROC curves analysis revealed sSIGLEC5 as a survival marker (cut-off ≤ 523.6 ng/mL). *In vitro* experiments illustrated how SIGLEC5 impaired CD8^+^ proliferation through binding to PSGL1. Blocking the SIGLEC5/PSGL1 axis reverted the latter effect. Mechanistically, SIGLEC5 overexpression was driven by HIF1α. Exogenous sSIGLEC5 accelerated death and magnified acute lung injury in mice models. Our data demonstrates how plasma sSIGLEC5 level on admission predicts death and stratifies patients with sepsis. This molecule exhibits the hallmarks of an IC ligand.

## INTRODUCTION

Sepsis is perceived as a leading cause of death in intensive care units (ICU) worldwide (1). Numerous therapeutic strategies to treat human sepsis other than antimicrobial and fluid resuscitation treatments have failed in clinical trials; hence, solid biomarkers of sepsis evolution remain to be identified. Though gold standards to define boundaries amongst sepsis and systemic inflammatory response syndrome (SIRS) still lack, an increasing number of studies have indicated that patients with systemic response to infection show most of the hallmarks of immunosuppression. In addition, in the complex dynamics of sepsis, two phases have been recognised: an early inflammatory phase and a late immunosuppressive phase (2, 3); however, these two phases can overlap (4). The early phase is characterised by a systemic inflammatory response, while during the later phase, immunosuppression and leukocyte deactivation increase risks of secondary infection, and a high mortality (3, 5). A number of evidences have emphasised how immunosuppression might contribute to increasing mortality risk in patients with sepsis (3–7). Along these lines, several authors have reported that monocytes and macrophages play an important position in orchestrating this immunosuppressive phenotype during sepsis (3, 8). T-cell exhaustion, through the crosstalk between immune checkpoint (IC) ligands expressed on monocytes and macrophages surfaces and their counterparts on T cells, is the main mechanism through which this immunosuppressive phenotype occurs (9). In this regard, we and other authors have demonstrated the role of Programmed cell death-protein 1 (PD-1) and its ligand (PD-L1) in this context (10–13).

Several teams have described that sialic acid-binding immunoglobulin-type lectins (SIGLECs) may play a significant role in the modulation of the immune response (14–17). These lectins present sialic acid recognition sites in an Ig-like-V-type domain. Interestingly, that domain is described in other proteins known, and postulated as, ICs or ICs ligands such as PD-1 and PDL1, being the structure through they interact (18). Among the fifteen members of SIGLEC family reported in humans, SIGLEC5 is the one that recognises a remarkable spectrum of sialic acid structures including α2–3, α2–6 and α2–8 linkage conformations, as well as the N-acetylneuraminic and N-glycosilneuraminic variants of sialic acid (19, 20). Until now, the physiological role of SIGLEC5 has been diverse and related to cell-cell interactions, pathogens recognition, clearance of apoptotic cells, negative cell signalling through soluble factors like Hsp70, and endocytosis of ligands (21, 22). Its soluble form, sSIGLEC5, has been associated with the highly sialylated P-selectin glycoprotein ligand-1 (PSGL1) molecule, showing a robust anti-inflammatory activity (23). Other authors have reported that PSGL1 has shown ability to induce CD8^+^ T-cell dysfunction acting as an IC regulator in a murine model of chronic virus infection (24).

Considering these assertions, we have applied a combination of *in vitro* and *in vivo* approaches to study the role of SIGLEC5 in the context of sepsis. We have analysed both its expression on blood monocytes and the circulating levels of the soluble isoform, in a wide variety of patients to establish its role as an IC ligand, as well as the molecular mechanisms involved. Furthermore, we have tested its usefulness as biomarker of severity in this pathology.

## RESULTS

### SIGLEC5 is a biomarker of prognosis in septic patients

A total of 425 patients with systemic inflammatory response criteria were recruited, and then classified as septic (44%), septic-shock (37.2%) and non-infectious SIRS (18.8%) according to their clinical parameters and severity (25, 26). They were followed-up for 60 days and then stratified into two groups: Survivors and *Exitus* (**Table 1**). As controls, healthy volunteers (HVs) and also patients suffering aneurysm or stroke were assessed (**Supplemental Table 1)**.

**Table 1.** Baseline characteristics of patients and healthy volunteers included in the study.

| **Characteristic \*** | **All Patients**<br>**(N=425)** | **Sepsis**<br>**(N=187)** | **Septic Shock**<br>**(N=158)** | **SIRS\*\***<br>**(N=80)** | **P-value†** |
| --- | --- | --- | --- | --- | --- |
| **Age - yr** | 67.4±16.2 | 67.2±17.2 | 69±15.1 | 65±15.7 |  |
| **Male sex – no. (%)** | 240(56.47) | 106(56.68) | 89(56.33) | 45(56.25) |  |
| **APACHE II Score** | 17.9± 7.6 | 17.3±6.2 | 22.2±8.3 | 13.3±4.4 | <0.0001 |
| **SOFA Score** | 5.3±3.2 | 4±2.4 | 7.5±3.3 | 4±2.1 | <0.0001 |
| **Lactate, nmol/L** | 3.28±3.21 | 2.95±2.39 | 4.49±4.28 | 1.66±0.76 | <0.0001 |
| **CRP, mg/L** | 73.37±105.39 | 75.65±103.17 | 70.62±107.95 | - |  |
| **PCT, ng/mL** | 25.73±51.73 | 20.52±51.54 | 31.94±51.35 | - | <0.0001 |

\*Data are presented as mean±SD, or number (%).
\*\*Non-infectious SIRS.
† P values were calculated by ANOVA test.
APACHE II: Acute Physiology and Chronic Health Evaluation II; CRP: C-reactive protein; INR, International Normalised Ratio; PCT: Procalcitonin; SIRS, Sitemic Inflammatory Response Syndrome; SOFA: Sequential Organ Failure Assessment.

Higher quantities of the soluble form of SIGLEC5, sSIGLEC5, were detected in plasma from all septic patients included in the study with respect to HVs (**Figure 1A**). Moreover, once septic patients were classified according to their severity in sepsis and septic-shock (26), we have learnt that levels of sSIGLEC5 also discriminate between these two groups, patients with SIRS and those with non-infectious diseases as aneurysm or stroke (**Figure 1B**). As septic patients were classified according to their evolution into survivors and *exitus*, levels of sSIGLEC5 were higher in the latter (**Figure 1C**). Eventually, to differentiate between both outcomes, the receiver-operating characteristic (ROC) analysis of plasmatic sSIGLEC5 levels was performed (**Figure 1D**). The area under the curve (AUC) was 0.702 (95% confidence interval [CI], 0.645 to 0.760; P<0.0001). The optimal cut-off, estimated by Youden index, was 523.6 ng/mL (**Figure 1E**), suggesting that patients with a circulating sSIGLEC5 level above 523.6 ng/mL exhibit a poor outcome. Collectively, these data illustrate that plasma concentrations of sSIGLEC5 could be postulated as an *exitus* predictor in septic patients.

**Figure 1.**
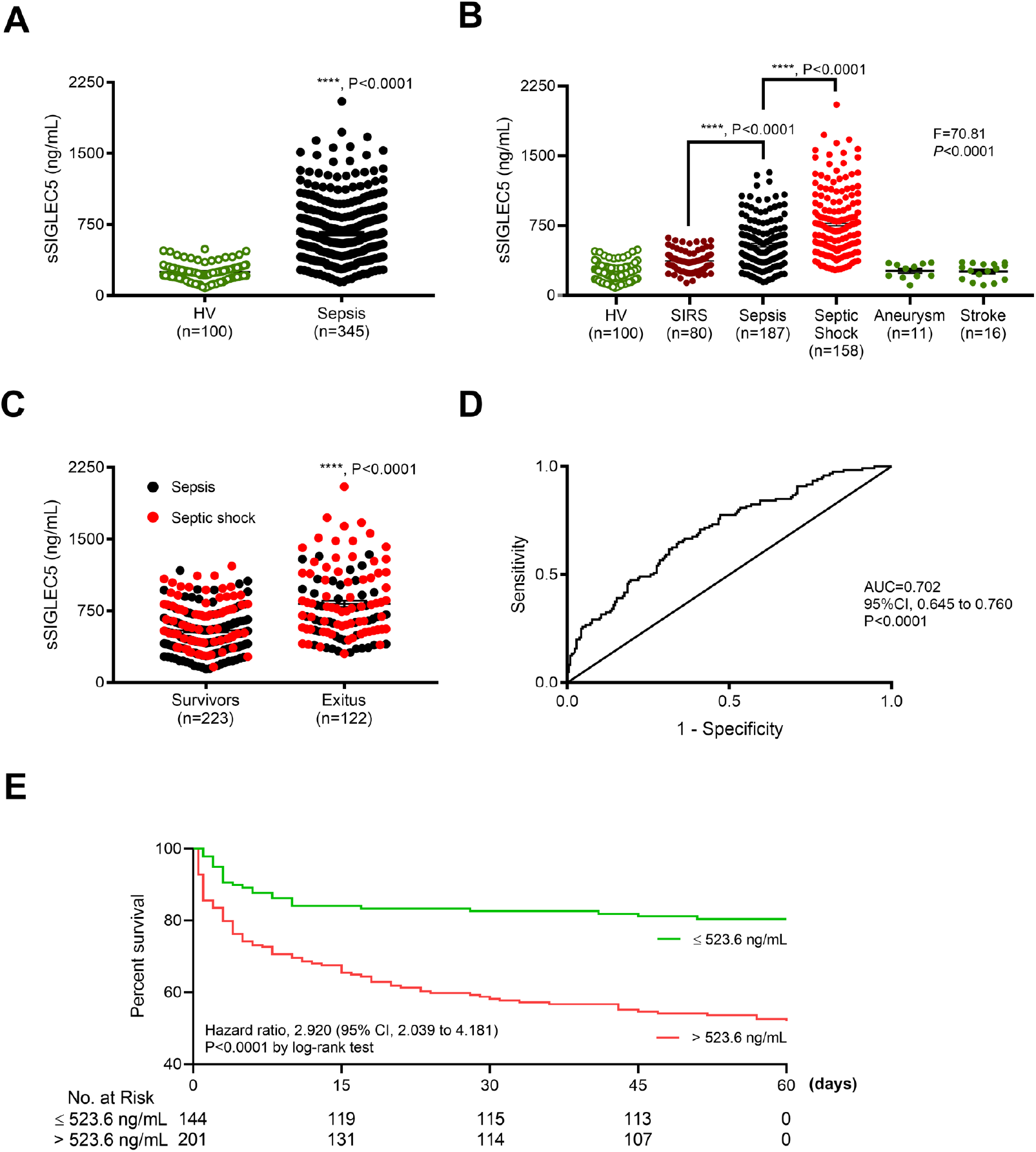
Soluble SIGLEC5 classified septic patients on admission.

**(A)** Soluble SIGLEC5 (sSIGLEC5) levels in plasma from HVs (n = 100) and patients with sepsis (n = 345). **(B)** sSIGLEC5 levels in plasma from HVs (n = 100) and patients with SIRS (n = 80), sepsis (n = 187), septic shock (n = 158), aneurysm (n = 11) and stroke (n = 16). **(C)** sSIGLEC5 levels in plasma from patients with Sepsis (black points) and Septic Shock (red points) classified according to their outcome after 60 days as Survivors (n = 223) and *Exitus* (n = 122). **(D)** Receiver-operating-characteristic (ROC) curve describing the predictive performance of plasmatic sSIGLEC5 concentration at sepsis diagnosis (black line; area under the curve [AUC] of 0.702 [95% CI, 0.645 to 0.760], *P*<0.0001), to identify which patients of the cohort (n = 345) would be dead within 60 days after diagnosis. **(E)** Patients were dichotomised according to the optimal cut-off, estimated by Youden index for plasmatic sSIGLEC5 concentration to be 523.6 ng/mL. Kaplan-Meier survival curves from diagnosis to day 60 according to baseline plasmatic sSIGLEC5 (_χ_2 = 27.13, P<0.0001) are shown. Statistical analysis was performed using unpaired t-test (**A, B** and **C**) or a one-way ANOVA followed by Tukey’s test (B). \*\*\*\**P*<0.0001.

### SIGLEC5 is produced by human circulating monocytes and HIF1α drives its expression

Since several authors have reported how monocytes and macrophages play an important role orchestrating the immunosuppressive phenotype during sepsis (3, 8), we evaluated SIGLEC5 expression on HVs’ monocytes challenged with LPS. As expected, they showed a patent upregulation of surface SIGLEC5 (**Figures 2, A and B**), as well as an increment of sSIGLEC5 in culture supernatants (**Figure 2C**). sSIGLEC5 appeared to be generated by metalloproteinase shedding, following a previous described mechanism (27). The presence of the pan-metalloproteinase inhibitor GM6001 (GM) increased and stabilised SIGLEC5 expression on monocytes’ surface and reduced sSIGLEC5 levels in supernatants (**Supplemental Figure 1**). Note that, SIGLEC5 expression was also evaluated on blood monocytes from random selected septic patients. **Figures 2, D and E** illustrate those differences between HVs and septic patients. We had previously reported the crucial role of HIF1α in the control of several key factors during sepsis, including the negative regulator of inflammation IRAK-M, the NLRP3 inflammasome activation and the IC ligand PD-L1 (10, 11, 28, 29). To explore whether *SIGLEC5* gene was a direct HIF1α target in humans, we searched for potential HIF1α binding sites in the proximal promoter region of human *SIGLEC5*, using bioinformatics approaches. As **Figure 3A** illustrates, we found four potential hypoxia response elements (HREs) containing the consensus sequence (5’-ACGTG-3’). Three of them were within *SIGLEC5* promoter and one in the middle of the transcript. Using Chromatin Immunoprecipitation (ChIP) assays, we demonstrated HIF1α binds to the three HREs identified in the promoter region of *SIGLEC5* in human monocytes, but not in the case of the HRE outside this promoter (**Figure 3B and Supplemental Figure 2A**). Subsequently, human HIF1α-transfected monocytes exhibited high expression of SIGLEC5 at both mRNA (**Supplemental Figure 2, B and C**) and protein levels (**Figures 3, C and D**). Eventually, SIGLEC5 expression on monocytes was reduced in the presence of PX-478 agent, a potent inhibitor of HIF1α (30) (**Figures 3, E and F** and **Supplemental Figure 3**).

**Figure 2.**
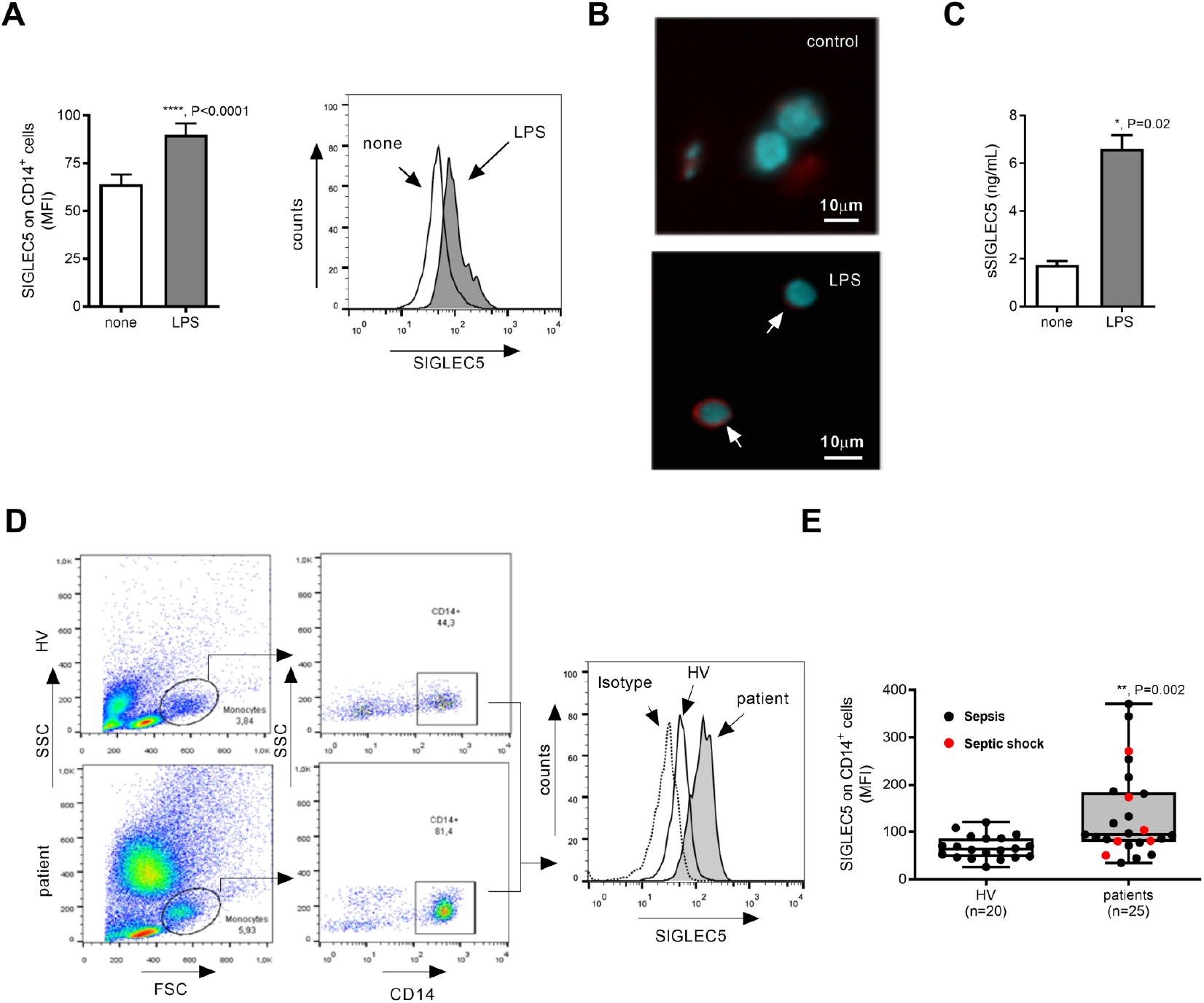
LPS induces SIGLEC5 expression on human monocytes and septic patients overexpress SIGLEC5. **(A)** SIGLEC5 expression on monocytes from HVs challenged or not with 10 ng/mL of LPS for 16 hours (n = 18) (left panel) and a representative histogram overlay (right panel). **(B)** Representative epifluorescence images (DAPI, blue; SIGLEC5, red) of SIGLEC5 expression on monocytes treated or not with 10 ng/mL of LPS for 16 hours. **(C)** sSIGLEC5 levels in supernatants of monocytes treated or not with 10 ng/mL of LPS for 16 hours (n = 3). **(D)** Flow cytometry gating strategy for CD14^+^ cells from both HVs and septic patient (left panels). Representative histogram overlay of SIGLEC5 expression on CD14^+^ cells from a HV and a septic patient (right panel). **(E)** Mean Fluorescence Intensity (MFI) of SIGLEC5 on CD14^+^cells from HV (empty box, n = 20) and patients (grey box, n = 25). Statistical analysis was performed using paired t-test (**A** and **C**) and an unpaired t test (**E**). \**P*<0.05*; ***P*<0.001; \*\*\*\**P*<0.0001.

**Figure 3.**
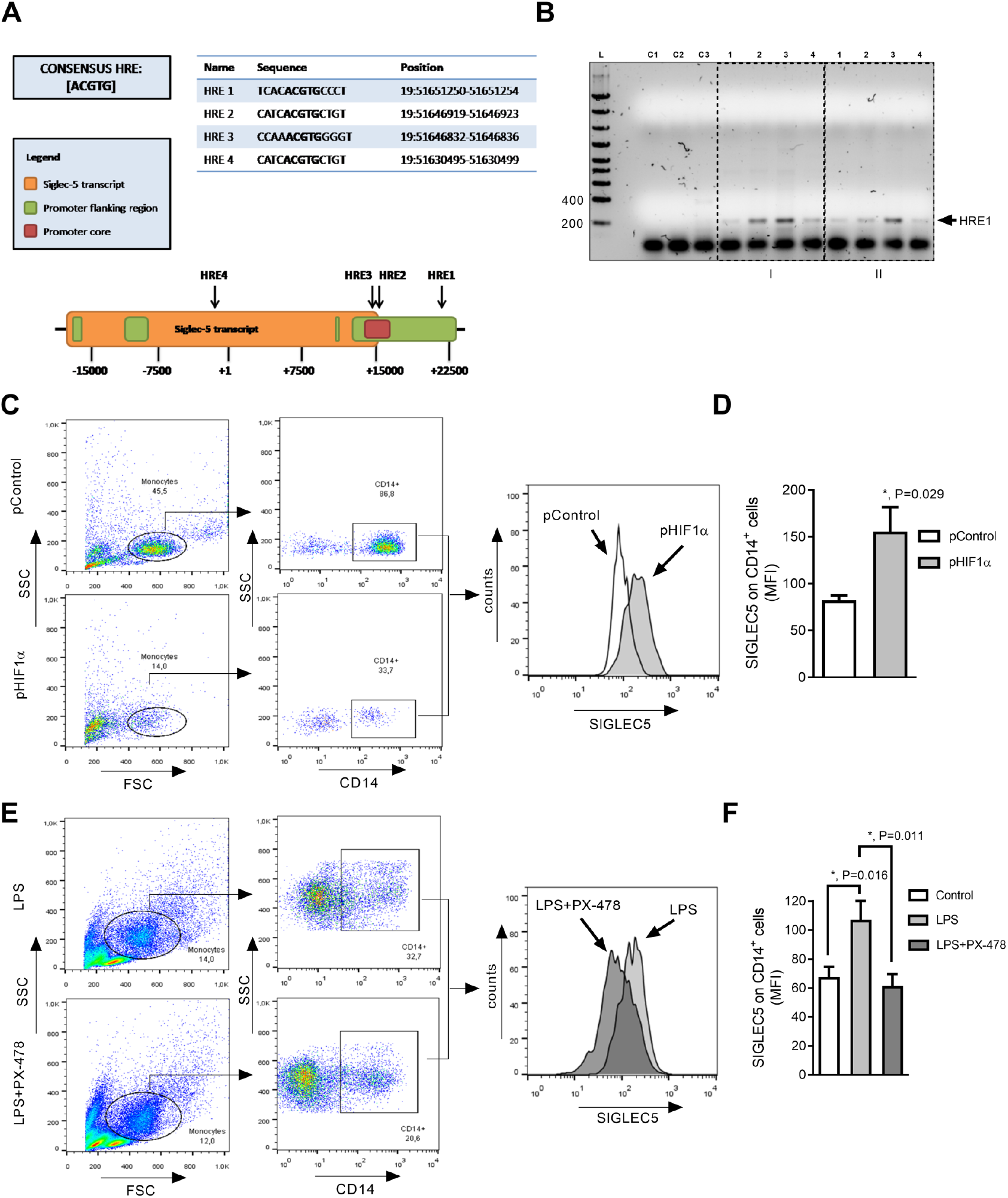
SIGLEC5 is expressed under Hypoxia Inducible Factor 1 alpha (HIF1α) control.

**(A)** Hypoxia Response Elements (HRE) in the human *SIGLEC5* sequence (Ensembl, ENSG00000105501) based on the consensus sequence [5’-ACGTG-3’]. Sequences and positions are shown. **(B)** Representative agarose gel from HRE1-SIGLEC5 analysis on two HV (I and II) of a Chromatin Immunoprecipitation (ChIP) assay conducted on: control monocytes (1), LPS-stimulated monocytes for 16 hours (2), HIF1α-transfected monocytes immunoprecipitated with either a HIF1α antibody (3) or an unspecific IgG antibody (4). HRE sites inside *SIGLEC5* promoter were analysed by PCR (see **Table S2**). Ladder (L), negative control using primers for HRE6-*PD-L1* region without the chromatin precipitate (C1), negative control using primers for HRE1-*SIGLEC5* region without the chromatin precipitate (C2), HIF1α-transfected monocytes immunoprecipitated with an antibody against HIF1α of a previously described HRE on *PD-L1* gen (C3). **(C)** Flow cytometry gating strategy of CD14^+^ cells nucleofected with pControl (left, upper panels) or pHIF1α (left, lower panels). Representative histogram overlay of SIGLEC5 expression on CD14^+^ cells nucleofected with pControl (empty) or pHIF1α (grey) (right panel). **(D)** MFI of SIGLEC5 on CD14^+^ cells after 16 hours of nucleofection with pControl (empty) or pHIF1α (grey) (n = 4). **(E)** Flow cytometry gating strategy of CD14^+^ cells pre-treated (left, lower panels) or not (left, upper panels) with a specific inhibitor (PX-478) of HIF1α for 3 hours, and then challenged with LPS for 16 hours. Representative histogram overlay of SIGLEC5 on CD14^+^ cells pre-treated (dark grey) or not (grey) with a specific inhibitor (PX-478) of HIF1α for 3 hours, and then challenged with LPS for 16 hours (right panel). **(F)** MFI of SIGLEC5 on CD14^+^ cells pre-treated (dark grey) or not (light grey) with a specific HIF1α inhibitor (PX-478) for 3 hours, and then challenged or not (empty box) with LPS for 16 hours (n = 7).). Statistical analysis was performed using paired t-test (**D** and **F**). \**P*<0.05.

### SIGLEC5 shows IC ligand hallmarks

We further analysed the potential ability of SIGLEC5 to modulate the immune response of T-cells. Thus, sSIGLEC5r reduced CD8^+^ cells proliferation *ex vivo* in a dose-dependent manner (**Figure 4A**, see gating strategy in **Supplemental Figure 4A**) and increased the apoptosis ratio in the same cells-subset (**Figure 4B**). Since this activity resembles ICs hallmarks, a multiple sequence alignment analysis of the Ig-like-V-type domain between SIGLEC5 and other proteins known and postulated as ICs or IC ligands, showed human SIGLEC5 shares a canonical Ig-like-V-type ICs’ domain in its primary structure, with identity and similarity ranges between 14 to 25% and between 22 to 38%, respectively (**Supplemental Figure 5)**. This homology is similar and even higher to that exhibited amongst those postulated ICs or IC ligands, like the B7 family proteins (18, 31, 32). Moreover, when LPS-stimulated monocytes from HVs were co-cultured with autologous lymphocytes, a reduced CD8^+^ cells ability for proliferating under stimulation by classical mitogen-lectin pokeweed (PWD) was observed (**Figure 4C)**. This effect was reverted when SIGLEC5 was blocked with either a SIGLEC5 antibody (**Figure 4C**) or when SIGLEC5 expression was siRNA blocked (**Figures 4, D and E**).

**Figure 4.**
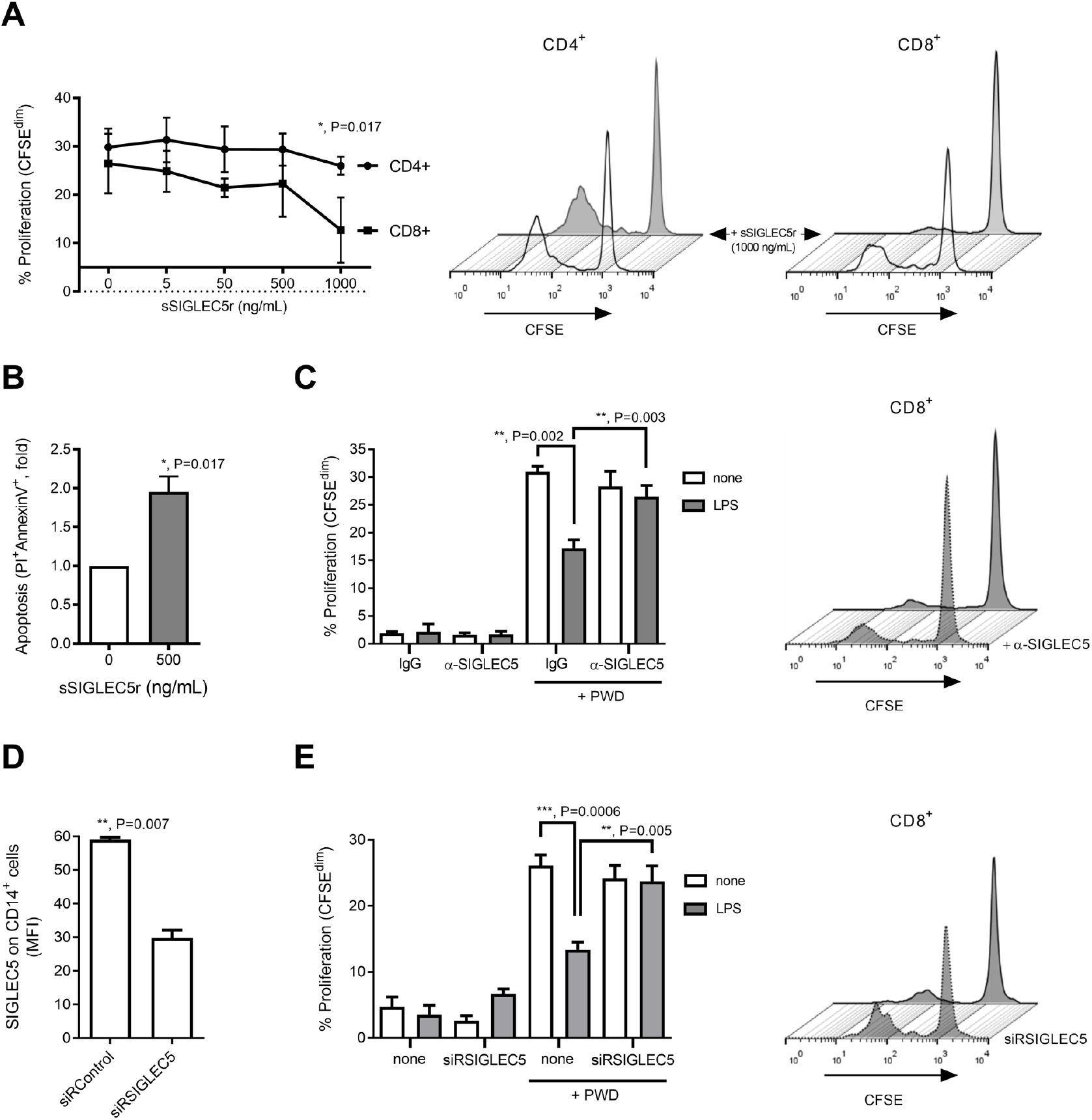
SIGLEC5 shows IC ligand hallmarks.

**(A)** Proliferation levels (CFSE^dim^) of PWD-stimulated CD4^+^ (sphere) and CD8^+^ (square) cells from HVs in presence of human recombinant sSIGLEC5r for 5 days (n = 5) (left panel). Representative histograms overlays of the proliferating CFSE^dim^ PWD-stimulated CD4^+^ (central panel) and CD8^+^ (right panel) cells from HVs in presence (grey) or not (empty) of human sSIGLEC5r for 5 days (see gating strategy in **Fig. S4A**). **(B)** Fold change of apoptotic (PI^+^/AnnexinV^+^) CD8^+^ cells isolated from HV and incubated or not with sSIGLEC5r for 24 hours (n = 5). **(C)** Proliferation levels (CFSE^dim^) of CD8^+^ cells from HVs, stimulated or not with PWD and co-cultured for 5 days with autologous monocytes pre-challenged (grey columns) or not (empty columns) with 10 ng/mL of LPS for 16 hours. In some indicated conditions, a blocking antibody against SIGLEC5 (α-SIGLEC5) or an unspecific IgG was added (n = 3, left panel). Representative histogram overlay of proliferation levels (CFSE^dim^) of PWD-stimulated CD8^+^ cells from HVs, and co-cultured for 5 days with autologous monocytes pre-challenged with 10 ng/mL of LPS for 16 hours, in presence or not of a blocking antibody against SIGLEC5, α-SIGLEC5 (right panel). **(D)** MFI of SIGLEC5 on CD14^+^ cells 16 hours after nucleofection with either siRSIGLEC5 or siRControl (n = 4). **(E)** Proliferation levels (CFSE^dim^) of CD8^+^ cells from HV, stimulated or not with PWD and co-cultured for 5 days with autologous monocytes prenucleofected or not with a specific siRNA against SIGLEC5 (siRSIGLEC5) and pre-challenged (grey columns) or not (empty columns) for 16 hours with 10 ng/mL of LPS (n = 4) (left panel). Representative histogram overlay of proliferation levels (CFSE^dim^) of PWD-stimulated CD8^+^ cells from HVs, and co-cultured for 5 days with autologous monocytes pre-challenged with 10 ng/mL of LPS for 16 hours and pre-nucleofected or not with siRSIGLEC5 (right panel). Statistical analysis was performed using paired t-test (**A-E**). \**P*<0.05; \*\**P*<0.01; \*\*\**P*<0.001.

We next studied how SIGLEC5 interacts with T-cells. **Figure 5A** shows the specific binding of the recombinant SIGLEC5-FC protein, sSIGLEC5r-FC, to CD8^+^ cells. This binding was reduced when lymphocytes were treated with neuraminidase (NM), a glycoside-hydrolase enzyme that cleaves the glycosidic linkage of neuraminic acids. Accordingly, NM also reduced the staining intensity of PSGL1 (a highly sialylated glycoprotein recently reported to interact with SIGLEC5 (23) on CD8^+^ cells (**Figure 5B**). Furthermore, a PSGL1 antibody reverted CD8^+^ cell proliferation when these cells were co-cultured with monocytes previously transfected to overexpress SIGLEC5 (**Figures 5, C and D**).

**Figure 5.**
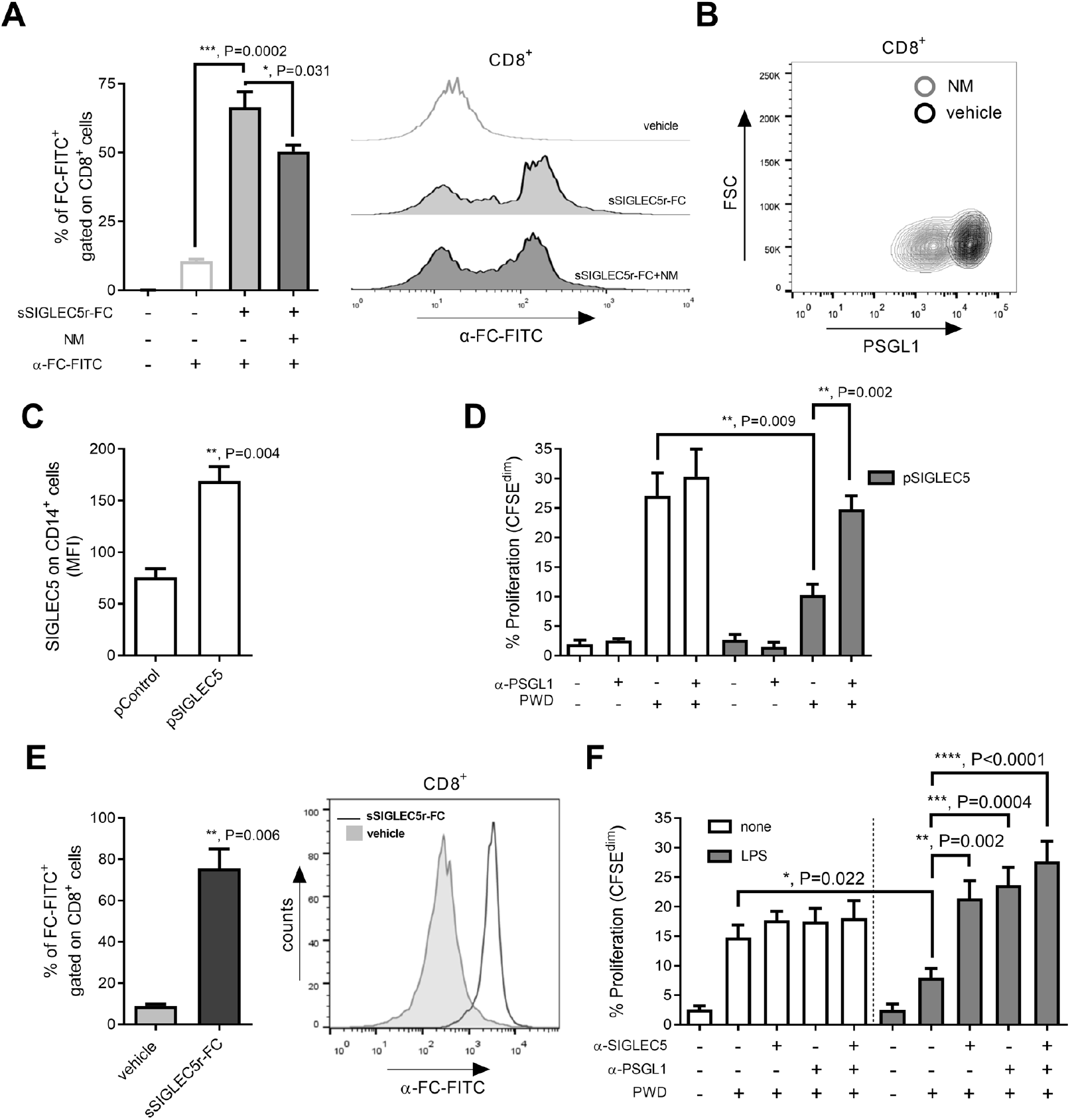
Blocking the SIGLEC5/PSGL1 axis restores T cells proliferation. **(A)** Quantification of sSIGLEC5r (sSIGLEC5r-FC) binding to CD8^+^ cells from HVs pre-treated (dark grey) or not (grey) with Neuraminidase (NM, 0.01 U/mL) for 24 hours (n = 6). The binding was revealed by an antibody (α-FC-FITC) and analysed by flow cytometry with an unspecific control using the antibody only (empty) (left panel). Representative histogram overlay of sSIGLEC5r-FC binding to CD8^+^ cells from HVs pre-treated (dark grey) or not (grey) with NM (0.01 U/mL)for 24 hours and unspecific control using the antibody α-FC-FITC only (empty) (right panel). **(B)** Representative contour plot of the PSGL1 expression on CD8^+^ cells pre-treated (grey) or not (dark grey) with NM (0.01 U/mL) for 24 hours. **(C)** MFI of SIGLEC5 expression on CD14^+^ cells after 16 hours of nucleofection with an expression vector of SIGLEC5 (pSIGLEC5) or an empty vector (pControl). **(D)** Proliferation levels (CFSE^dim^) of CD8^+^ cells from HVs in presence or not of PWD and co-cultured for 5 days with autologous monocytes pre-nucleofected with an expression vector of SIGLEC5 (pSIGLEC5, grey columns) or an empty vector (pControl, empty columns) (n = 6). In some conditions a blocking antibody against PSGL1 (α-PSGL1) was added. **(E)** sSIGLEC5r-FC binding quantification on CD8^+^ cells from patients with sepsis (n = 4) (left panel) and a representative histogram overlay (right panel). See gating strategy and **Figure S4B. (F)** Proliferation levels (CFSE^dim^) of CD8^+^ cells from patients with sepsis in presence or not of PWD and co-cultured for 5 days with autologous monocytes pre-challenged with 10 ng/mL of LPS for 16 hours in presence of anti-SIGLEC5 blocking antibody and/or anti-PSGL1 blocking antibody (n = 6). Gating strategy and a representative histogram overlay are shown in **Figure S4C**. Statistical analysis was performed using paired t-test (**A, C, D, E** and **F**). \**P*<0.05; \*\**P*<0.01; \*\*\**P*<0.001; \*\*\*\**P*<0.0001.

As expected, SIGLEC5 not only binds CD8^+^cells from HVs but also those from septic patients (**Figure 5E**, see gating strategy in **Supplemental Figure 4B**). Therefore, CD8^+^lymphocytes from septic patients showed an impaired proliferation, which was reverted in the presence of either anti-SIGLEC5 or anti-PSGL1 antibodies (**Figure 5F** and **Supplemental Figure 4C**). Despite CD4^+^ cells bound sSIGLEC5-FC their proliferation was not affected (**Supplemental Figure 6**).

### SIGLEC5 reduces viability in mice models

To better understand the role of SIGLEC5 in immune response, two *in vivo* mice models were developed. Firstly, a caecal ligation and puncture (CLP) protocol was performed on mice infused with sSIGLEC5r. Mortality in sSIGLEC5r-CLP-treated mice was accelerated respect to CLP-shams (_χ_2 = 7.591, P = 0.006, Hazard ratio CLP+sSIGLEC5r *vs*. CLP of 8.302; 95% CI, 1.842 to 37.420, **Figure 6A**). In line, **Figure 6C** illustrates how those mice treated with LPS+sSIGLEC5r died earlier than those challenged only with LPS (_χ_2 = 5.432, P = 0.019, Hazard ratio LPS+sSIGLEC5r *vs*. LPS of 7.123; 95% CI, 1.366 to 37.13). No differences were found between the controls and sSIGLEC5r-treated mice. Additionally, when CLP protocol was performed using lymphocytes-deficient mice (*Rag1*^−/-^), sSIGLEC5r infusion did not influence survival rate (_χ_2 = 0.064, P n.s., Hazard ratio CLP+sSIGLEC5r *vs*. CLP of 0.83; 95% CI, 0.161 to 3.582, **Figure 6E**). These data suggested that sSIGLEC5 effects on mortality are related to lymphocytes.

**Figure 6.**
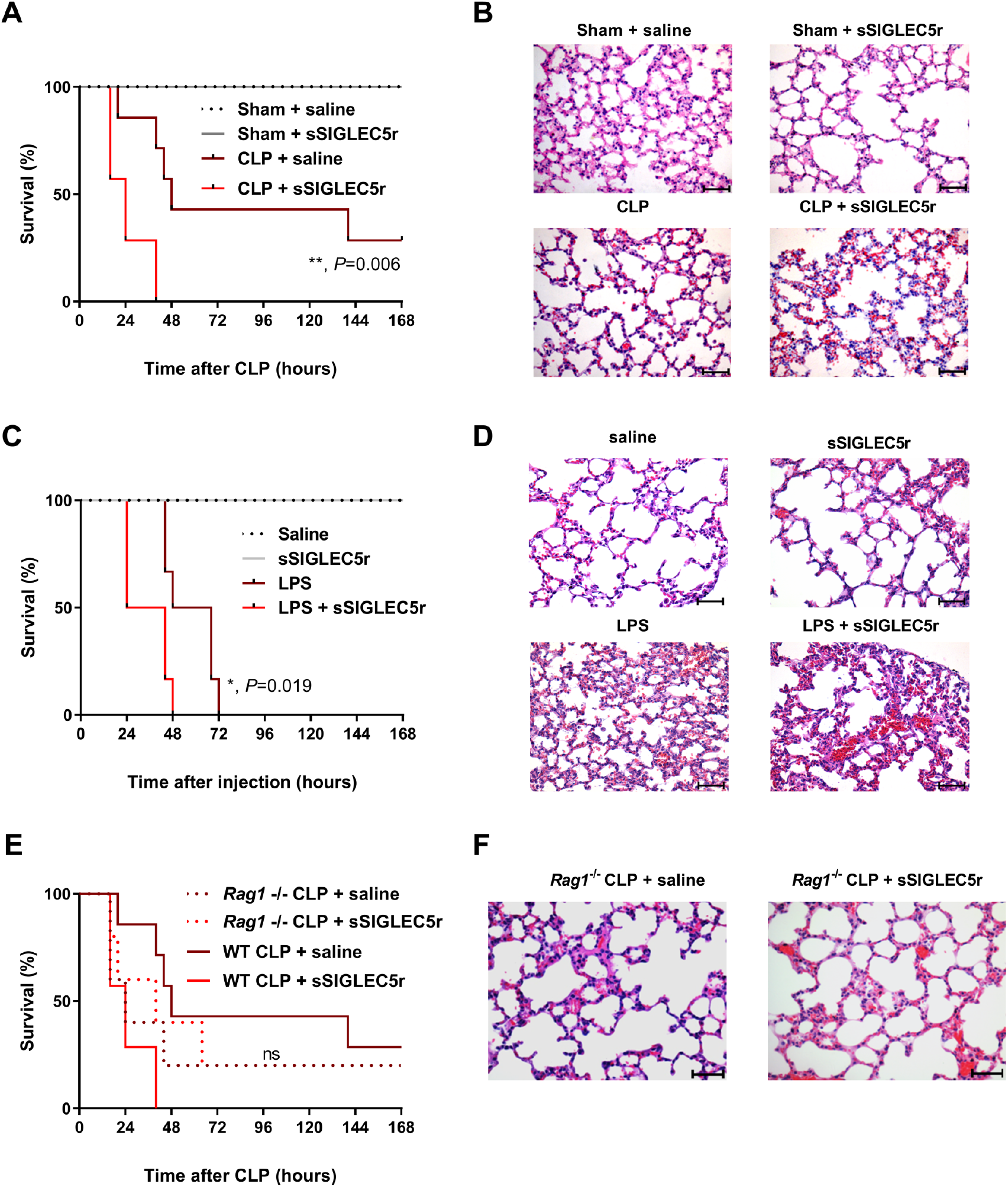
SIGLEC5 accelerates death in animal models of sepsis. **(A)** Kaplan-Meier estimation of progression-free survival from CLP-mice injected or not with sSIGLEC5r (_χ_2 = 33.20, *P*<0.0001 for all four groups comparison; _χ_2 = 7.591, \*\**P* = 0.006, hazard ratio for death [HR] of 8.302; 95% CI, 1.842 to 37.42 for CLP plus sSIGLEC5r *vs*. CLP groups comparison). **(B)** A set of representative images of Haematoxylin/Eosin stained lung sections from mice of the indicated experimental group at 400x magnification; scale bar 50 ¼m are shown. **(C)** Kaplan-Meier estimates of progression-free survival from mice injected with sSIGLEC5r and/or LPS (_χ_2 = 32.07, *P*<0.0001 for all four groups comparison; _χ_2 = 5.432, *P* = 0.019, Hazard ratio for death [HR] of 7.123; 95% CI, 1.366 to 37.13 for LPS plus sSIGLEC5r *vs*. LPS groups comparison). **(D)** A set of representative images of Haematoxylin/Eosin stained lung sections from mice of the indicated experimental group at 400x magnification; scale bar 50 ¼m are shown. **(E)** Kaplan-Meier estimation of progression-free survival from CLP-WT or CLP-*Rag1^−/−^* mice injected or not with sSIGLEC5r (_χ_2 = 0.064, *P* n.s., Hazard ratio [HR] *Rag1^−/−^* CLP vs. *Rag1^−/−^* CLP + sSIGLEC5 of 0.83, 95% CI, 0.161 to 3.582). **(F)** A set of representative images of Haematoxylin/Eosin stained lung sections from mice of the indicated experimental group at 400× magnification; scale bar 50 ¼m are shown.

To all models, haematoxylin/eosin staining corroborated a substantial lung injury in CLP or LPS+sSIGLEC5r groups of mice (**Figures 6, B, D and F and Supplemental Figures 7, A and B**). Besides, apoptosis rate of mice CD8a^+^ splenocytes, isolated and cultured for 24 hours, was higher in those cells coming from mice treated with LPS+sSIGLEC5r than with LPS alone (**Supplemental Figure 7C**).

## DISCUSSION

An increasing number of studies highlight the importance of immunosuppression and the alternative non-inflammatory activation of the innate immune system during sepsis evolution. Whilst over-inflammation can be controlled with steroids and antibiotics, deaths from sepsis reflect a host immunosuppression that implies a high risk to nosocomial infection (33, 34). In fact, numerous anti-inflammatory therapies have been developed against the pro-inflammatory phase of sepsis. However, all these therapies, including anti-endotoxin, anti-TNFα, anti-IL1, and Toll-like receptor inhibitors (35–37), have failed in some clinical trial phases. In contrast, the study of the immunosuppressive mechanisms in septic patients is gaining ground in the research into this disease. In this regard, ICs and their ligands appear to play a crucial role in the prognosis and treatment of septic patients.

Early diagnosis and stratification of sepsis severity is crucial to determine the fate of patients in the ICU (38). Robustness of scores to identify patients at low risk, which may require deescalation of the intensity of care, will improve actual efforts on those with poor prognoses at the ICU. We have reported the increment of SIGLEC5 expression on circulating monocytes during sepsis, which is reflected in its soluble form levels in plasma from septic patients but not in patients with SIRS, aneurysm, stroke or HVs. Note that sSIGLEC5 could be generated by a shedding event involving metalloproteinases. Plasma levels of sSIGLEC5 aid distinguish between sepsis and septic-shock and it could additionally serve as a biomarker of sepsis evolution, being higher in patients with poor prognoses. Along these lines, ROC analysis showed a robust AUC for sSIGLEC5 as an exitus predictor in sepsis. Thereby, the cut-off value calculated based on the Youden index, revealed that plasma levels over 523.6 ng/mL of sSIGLEC5 on admission and before any treatment indicated a poor prognosis. This data may constitute a feasible score to incorporate into routine analysis of patients achieving sepsis criteria upon arrival at the ICU. Its importance is reinforced by data observed in mice models, which corroborate that exogenous sSIGLEC5r increases mortality in both CLP and mice treated with LPS from Escherichia coli. Interestingly, lymphocytes-deficient mice did not exhibit different mortality rate after CLP when they were treated with exogenous sSIGLEC5r. Thus, SIGLEC5 not only could be a biomarker of severity in sepsis but also acting as an IC ligand (39). In line, SIGLEC5 structure exhibited similarities with well-known ICs and induced patent effects on CD8+ cells exhaustion. Furthermore, several authors have indicated that the number of these effector cytotoxic cells decreased in patients with septic-shock on admission (40). It is very well established that composition of the naïve pathogen-specific CD8^+^ cells repertoire is important in both the clearance of infection and the generation of memory CD8^+^ cells in response to pathogens (41). Thus, an impaired CD8^+^ proliferation together with their low viability compromise the evolution of septic patients, increasing their risk of suffering from secondary infections and death.

Our findings also indicated a manifest binding of SIGLEC5 to CD8^+^cells, in a PSGL1-dependent manner. PSGL1 had been described as an IC receptor that promotes CD8^+^ T-cell exhaustion in murine model of chronic virus infection (24). Nevertheless, its ligand was unknown until lately(42). Our findings postulate SIGLEC5 as a ligand of PSGL1 in a sialic acid-dependent binding. Moreover, blockade of SIGLEC5/PSGL1 crosstalk, reverted the impaired CD8^+^ proliferation and blocked their apoptosis, suggesting this could be a new pharmacological target in the treatment of septic patients.

We had demonstrated the involvement of HIF1α in the regulation of both the innate and the adaptive response during sepsis, including the control of the transcription of a set of genes, such as IRAKM, VEGFA, and MMP (10, 11, 28). Now, three HREs are corroborated in human SIGLEC5 promoter. Mechanistically, our data widens the known HIF1α control over the expression of SIGLEC5 on monocytes and the induction of T-cells exhaustion during sepsis. We, along with others, had also demonstrated PD-L1 expression on circulating monocytes during sepsis (10, 11). However, despite blockage of PD-L1/PD1 interaction using an anti-PD1 antibody led to a revolutionary treatment for many types of cancers (43), no clear results are available in sepsis. Data from mouse models have indicated that administration of anti-PD-L1/PD1 antibodies prevents lymphocyte depletion (44) and improves survival in Candida albicans sepsis (45). In addition, at least one clinical trial has tested the inhibition of PD-L1/PD1 crosstalk (NCT02576457): BMS-936559 inhibitor was well tolerated, with no evidence of hypercytokinemia or cytokine storm, and some restoration of the immune status was reported (46), although data are not conclusive. Herein, we have reported another IC ligand with a significant role in the prognosis of septic patients. Moreover, the expression of SIGLEC5 on circulating monocytes could provide a negative signal for migration to the infected sites and their interaction with local lymphocytes (23). In terms of a clinical application, one of the first questions to emerge would be for which septic patients to apply a potential SIGLEC5/PSGL1 blocking strategy and at what time window. According to our findings, measuring levels of sSIGLEC5 on admission is required.

## METHODS

### Healthy volunteers and patients’ samples

Patients older than 18 years meeting the diagnostic criteria for sepsis according to the Third International Consensus Definitions for Sepsis and Septic-Shock (26) (Sepsis-3) were enrolled in this study upon arrival to Emergencies (Table 1). Sepsis picture was primarily identified by SOFA (tachycardia, altered mentation and hypotension) and then thoroughly corroborated following International Sepsis Definitions Conference criteria (25) and classified according to their severity in sepsis and septic shock (operationally defined as requiring vasopressor therapy to maintain a mean arterial blood pressure of > 65 mmHg and an increased plasma lactate level of > 2 mmol/L). Patients were followed-up for 60 days and classified according to their outcome into Survivors or Exitus. Blood samples from patients were taken within the first hour after admission and peripheral blood mononuclear cells (PBMCs) and plasma were isolated by standardised procedures (47). Biobank samples and data from the patients were provided by the Sepsis Bank of Vall d’Hebron University Hospital Biobank and Virgen de la Arrixaca University Clinical Hospital Biobank (Spanish National Biobanks Network).

As controls, healthy volunteers (n = 100) were recruited from the Blood Donor Services of La Paz University Hospital and patients with aneurysm (n = 11) and stroke (n = 16) were also enrolled from The Emergency Department and Department of Neurology of La Paz University Hospital, respectively (see Supplemental Table 1). All of them were free of pathogen colonisation.

### Mouse strains

Wild-type female C57BL/6 mice (6–9 weeks old) were purchased from Charles River Laboratories (France). Rag −/− female C57BL/6 mice (8–9 weeks old) were a gift from Carlos del Fresno (CNIC, Madrid, Spain). Mice were properly housed in temperature and light-regulated rooms (12:12 hours light/dark cycle, 21–24ºC) with food and water ad libitum. All mice were randomly assigned to experimental groups.

### Cell culture

PBMCs and plasma from healthy volunteers and patients were isolated by Ficoll-Plus (GE Healthcare Bio-Sciences) gradient. PBMCs were cultured in RPMI 1640 media containing 10% foetal bovine serum (PBS), 25 mM HEPES, L-glutamine and 1% Penicillin and Streptomycin Mix (Gibco). Monocytes were enriched by: (1) adherence by plating PBMCs in FBS-free RPMI 1640 media with antibiotics for 1 hour, and then non-adherent cells were removed by washing with Phosphate Buffered Saline (PBS) three times; or (2) negative isolation using magnetic beads of the Monocyte Isolation Kit (Mintenyi Biotec) according the manufacturer’s instructions. The purity of the monocyte cultures was tested by CD14 labelling and FACS analysis (average 89% and > 95% of CD14+ cells, respectively). Enriched Monocytes were cultured in RPMI 1640 media containing 10% foetal bovine serum (PBS), 25 mM HEPES, L-glutamine and 1% Penicillin and Streptomycin Mix (Gibco). Splenocytes were obtained from spleens of WT female C57BL/6J mice, cultured in RPMI 1640 media containing 10% foetal bovine serum (PBS), 25 mM HEPES, L-glutamine and 1% Penicillin and Streptomycin Mix (Gibco).

### ELISA protocol

Concentrations of sSIGLEC5 in supernatants of human monocyte cultures, and plasma samples, were determined using a commercially available ELISA kit (Sigma-Aldrich), following the manufacturer’s instructions.

### Antibodies and flow cytometry analysis

FACS analysis were developed using specific human antibodies (Abs) to the following surface molecules: CD8-Allophycocianin (APC), CD14-APC, HLA-DR-Fluorescein-isothiocyanate (FITC) (all three from ImmunoStep), CD3-Brilliant Violet (BV)-786, CD4-Peridininchlorophyll-A protein (PerCP), CD8-BV510, CD14-BV395, PSGL1-Phycoerythrin (PE) (all five from BD Biosciences) and SIGLEC5-PE (Miltenyi Biotec), as well as specific mouse CD8a-APC (Miltenyi Biotec).

Cells were stained with proper Abs for 30 minutes at 4ºC in the dark, and washed with Phosphate Buffer Saline (PBS) twice. Matched isotype antibodies were used as negative controls. For all assays, samples were run in FACS Calibur or FACS Celesta (BD Biosciences) flow cytometers, and data were analysed with FlowJo (TreeStar) v. X.07 software or BD FACSDiva v8, respectively.

### *In vitro* endotoxemia model

Monocytes (2·105 cells per well) were stimulated or not with 10 ng/mL Lipopolysaccharide (LPS, from Escherichia Coli, Sigma-Aldrich) for 16 hours in flat bottom 24-wells plates, in a final volume of 0.5 mL. For the time-course of extracellular SIGLEC5 expression, monocytes were treated or not with 10 ng/mL LPS, for periods ranging from 3 to 48 hours. For the matrix metalloproteinase (MMP) and ADAM-family inhibition, the specific inhibitor GM6001 (Merck Millipore) was added 6 hours after the LPS challenge, and cultures were maintained for a maximum of 48 hours. For HIF1α inhibition, the potent inhibitor PX-478 (Cayman Chemical) was added for 3 hours, and then challenged or not with LPS (10 ng/mL) for another 16 hours.

### Epifluorescence microscopy

For assessing the previously described SIGLEC5 protein expression on monocytes with or without LPS treatment, cells were fixed with 4% PFA for 30 minutes at room temperature (RT) and washed with PBS. Cells were then examined with epifluorescence and images were taken using Leica DMI6000B microscope.

### *In silico* analysis of genomic and protein sequences

Bioinformatics analysis, such as protein and genomic structure of SIGLEC5, were performed using the ENSEMBL, Uniprot and NextProt online tools. The proximal promoter region and flanking promoters of the human SIGLEC5 were localised using the ENSEMBL online tool, and the Hypoxia Response Elements (HREs) using a Python Script with the consensus site to the binding of the HIF1α factor (5’-ACGTG-3’). The multiple sequence alignments were performed with ClustalW (NCBI). To check the sequence identity and similarity of SIGLEC5 with the other proteins, the EMBOSS-Needle online tool was used.

### Monocytes nucleofection procedure

Magnetic negative sorted monocytes by Pan Monocyte Isolation Kit (Miltenyi Biotec) were used for HIF1α and SIGLEC5 overexpression and knockdown assays (see Cell Culture above) by nucleofection as previously described (48), using an Alexa Nucleofector (Lonza Group). HIF1α overexpression was done with an expression vector, which was a kind gift from Dr del Peso (Institute for Biomedical Research Alberto Sols). SIGLEC5 overexpression and knockdown, were done with an expression vector (pUNO1-hSIGLEC5, InvivoGen) and siRNA (s16725, ThermoFisher Scientific), respectively.

### Chromatin Immunoprecipitation (ChIP) Assay

Human/Mouse HIF1α ExactaChIP Chromatin IP Kit (R&D Systems) was used to perform ChIP on lysates from isolated human blood monocytes, previously nucleofected or not with a HIF1α overexpression vector (48). Resultant DNA was isolated using either an unspecific IgG antibody or a specific antibody against HIF1α, in order to perform the immunoprecipitation according to the manufacturer’s protocol.

The products (Hypoxia Response Elements [HRE] sites) were amplified with primers shown in Supplemental Table 3, using the NZY Jaq 2X Colourless Master Mix, in an Eppendorf AG Mastercycler Thermocycler. The following program was used: a first denaturation step at 94°C for 5 minutes, 40 cycles of denaturation at 94°C for 1 minute (primer annealing at 56°C for 50 seconds) with primer extension at 72°C for 45 seconds, and a final extension at 72°C for 5 min after completion of the cycling steps.

### RNA isolation and RT-qPCR

Adherent monocytes were washed twice with PBS and the RNA was extracted using the High Pure RNA Isolation Kit (Roche Diagnostics). The RNA concentration of each sample was measured using a NanoDrop 2000 (Thermo Fisher Scientific). cDNA was synthesised from 0.25μg total RNA using the High Capacity cDNA Reverse Transcription Kit (Applied Biosystems). RT-qPCRs were performed using the QuantiMix Easy SYG Kit (Biotools) according to the manufacturer’s instructions. Gene expression levels were analysed using the LightCycler system (Roche Diagnostics). Reactions were run in triplicate and expression level of β-ACTIN housekeeping was used as internal standard to normalise data. The cDNA copy number of each gene of interest was determined using a 7-point standard curve. All the primers were synthesised by Eurofins Genomics. Specific primers for each gene are shown in **Supplemental Table 2**.

### Lymphocytes proliferation assays

Carboxyfluorescein succinimidyl ester (CFSE) was purchased from ThermoFisher Scientific, and used following the manufacturer’s protocol to assess T cells lymphocytes proliferation. Isolated or nucleofected monocytes (104 cells per well) were seeded into round bottom 96-wells plates, and then stimulated or not with LPS (10 ng/mL) for 16 hours. Afterwards, monocytes were washed once with PBS and autologous CFSE-labelled T cells were added to a ratio 1:5 (monocyte:lymphocyte) in fresh complete RPMI 1640 medium. Cells were then stimulated or not with Pokeweed (PWD, 2.5 μg/mL), and treated or not with fully mouse anti-PSGL-1 antibody (1 μg/mL) or fully human anti-SIGLEC5 antibody (500 ng/mL).

### Apoptosis Assay

PBMCs (2·105 cells per well) from HVs, as well as mice splenocytes from in vivo endotoxemia model (2·105 cells per well), were cultured with or without sSIGLEC5r (500 ng/mL) for 24 hours, and then stained with AnnexinV-FITC and Propidium Iodide (PI), for measuring apoptosis by flow cytometry, following manufacturers’ recommendations (ImmunoStep).

### Binding Assay protocol

A polyclonal antibody against human IgG-Fc FITC-labelled (Abcam), hereinafter α-Fc-FITC, was purchased. A recombinant human SIGLEC5 protein, with a crystallisable fragment (Fc) region of antibody domain (R&D Sytems), hereinafter sSIGLEC5r, was purchased.

Binding of sSIGLEC5r was performed as previously reported (49). Briefly, PBMCs from HVs or septic patients, treated or not with neuraminidase (NM, 0.01 U/mL for 24 hours), were washed twice with 10 mL of ice-cold Ligand Binding Buffer (LBB, PBS with 1% BSA, 0.05% Sodium Azide and 0.1 mM CaCl2·2H2O), fixed using 4% paraformaldehyde (PFA), and 106 cells resuspended in 1 mL LBB. Afterwards, fixed PBMCs were washed twice with ice-cold LBB and incubated with the recombinant protein sSIGLEC5r (500 ng in 100 μL of LBB) or LBB alone for 1 hour and hand-mixed each 15 min on ice. Next, sSIGLE5r binding was detected using α-Fc-FITC (1 μL in 100 μL of LBB). Possible Fc-Receptors were blocked using a Fc-blocking (ImmunoStep) for 30 minutes before incubation with the recombinant protein. PBMCs incubated only with α-Fc-FITC were used as the negative control.

### LPS-induced endotoxemia *in vivo* model

C57BL/6 (6 weeks old; purchased to Instituto de Investigaciones Biomédicas “Albert Sols”, Madrid, Spain) female mice were administered a tail vein injection with either LPS (20 mg/kg), sSIGLEC5r (0.8 mg/kg) or both LPS (20 mg/kg) plus sSIGLEC5r (0.8 mg/kg) using sterile saline solution (0.90% w/v of NaCl) as vehicle. Mice were followed-up for survival and post-mortem samples were collected.

### Caecal Ligation and Puncture (CLP) *in vivo* model

C57BL/6 wild type (8–9 weeks old) female mice and Rag-1−/− C57BL/6 (8–9 weeks old; purchased to Centro Nacional de Investigaciones Cardiovasculares, Madrid, Spain) were modelled. Caecal ligation and puncture protocol was used to induce polymicrobial sepsis as described (50). Briefly, a 2-cm midline incision was performed to exteriorise the caecum, before being ligated with a 3.0-silk suture at 1 cm of its base, then punctured once with a 25-gauge needle and a small drop of caecal content from the perforation site extruded. Caecum was then returned to the peritoneal cavity and the abdominal incision closed with 4.0-silk sutures. Mice were administered or not a retro-orbital injection of sSIGLEC5r (0.8 mg/kg) using sterile saline solution (0.90% w/v of NaCl) as vehicle. Mice were followed-up for survival and post-mortem samples were collected.

### Histological analysis

Following sacrifice, lungs from mice were fixed in 4% paraformaldehyde embedded in paraffin and cut into 5μm sections in a microtome (RM2255, Lefor histopathological analysis. Leica Biosystems, Wetzlar, Germany). Sections were stained with haematoxylin and eosin (H&E) and images were captured using light microscopy (BX41, Olympus Optical Co Ltd, Tokyo, Japan). Briefly, the acute lung injury score of mice, on a scale of 0 (optimal) to 12 (severe), were estimated in accordance with combined assessments of alveolar congestion, haemorrhage, fibrin and infiltrates as previously described (51). Four randomly selected fields from each slide were analysed by three independent observers.

### Statistics

Data are presented as numbers and percentages, means and standard deviations. Accordingly, differences among the groups were evaluated with the use of Chi-square test, Student’s t-test or an analysis of variance (ANOVA) model, followed by the Tukey-Kramer test when findings with the ANOVA models were significant.

For patients with sepsis, Receiver-Operating Characteristic (ROC) curve analysis and its area under the curve (AUC) were used to determine whether the sSIGLEC5 levels could be used as a predictor of mortality in patients with sepsis. The area under the ROC curve measures how well the model discriminates between Survival and Exitus groups of patients. We also indicated sensitivity and specificity and the optimal threshold (in terms of maximum sensitivity and specificity) in the primary study cohort were determined on by the Youden Index. Considering sSIGLEC5 levels, we used the Kaplan-Meier method to estimate survival rate over time (60 days) and hazard ratio.

*P* values of less than 0.05 were considered to indicate statistical significance. All P-values are two-sided, and the 95% Confidence Intervals (95% CI) are also presented. Statistical analyses were conducted using Prism 6.0 (GraphPad), SPSS version 23 (IBM) software. Youden Index was calculated with MedCalc statistical software.

### Study approval

All patients and controls signed informed consent and the data were treated according to recommended criteria of confidentiality, following the ethical guidelines of the 1975 Declaration of Helsinki. The study was approval by the local Ethics Committee (La Paz University Hospital, Madrid, PI-3761). All mice experiments were designed according the guidelines and performed following approval by the Ethics Committee for Animal Experimentation of the Institutional Animal Care and Use Committee at Biomedical Research Institute (IIB, PI-2599).

## Data Availability

All data referred to in the manuscript are available.

## AUTHORS’ CONTRIBUTIONS

ELC designed the study. ELC and LAA wrote the manuscript. RLR and JAO performed the main experiments and discussed the results. PP, CG-P, JCR-R, RF, JJ G-L, AF, AMQ and ChME recruited the septic patients and followed-up them. MGF and EAL recruited patients with stroke, CCZ contributed samples from patients with aneurysms. ELG, KMH, VeT, ViT, EM, JV and RPD performed specific in vitro experimental assays. LAA, CdF, ViT, VeT, JAO, PS and RLR performed in vivo assays. PGC, RLR and JAO performed statistical analyses. RLR, JAO, RPD, PP and CdF performed a critical review of the manuscript. All authors read and agreed to submit the manuscript for publication.

## ACKNOWLEDGEMENTS

We acknowledge to Sepsis Bank of Vall d’Hebron University Hospital Biobank (Barcelona, Spain) and Virgen de la Arrixaca University Clinical Hospital (“Biobanco en Red de la Región de Murcia” Murcia, Spain) for plasma samples. This work was supported by grants from Instituto de Salud Carlos III (ISCIII) and “Fondos FEDER” to ELC (PIE 15/00065, PI 18/00148, PI 14/01234) and to PP (20859/PI/18) and received funding from the European Union’s Horizon 2020 research and innovation program under the Marie Sklodowaska-Curie grant agreement to KMH (No. 713673; “laCaixa”). CdF was supported by AECC Foundation (INVES192DELF).

The Vall d’Hebron University Hospital and Vall d’Hebron Research Institute were supported by Plan Nacional de I+D+i 2013–2016 and Instituto de Salud Carlos III and Spanish Network for Research in Infectious Diseases (REIPI RD16/0016/0003)—cofinanced by European Development Regional Fund “A way to achieve Europe”, and by the European Union’s Horizon 2020 Research and Innovation Program. Authors thank Emilio Llanos for his technical assistance.

## Supplemental Data

### Supplementary Figures

**Supplemental Figure 1.**
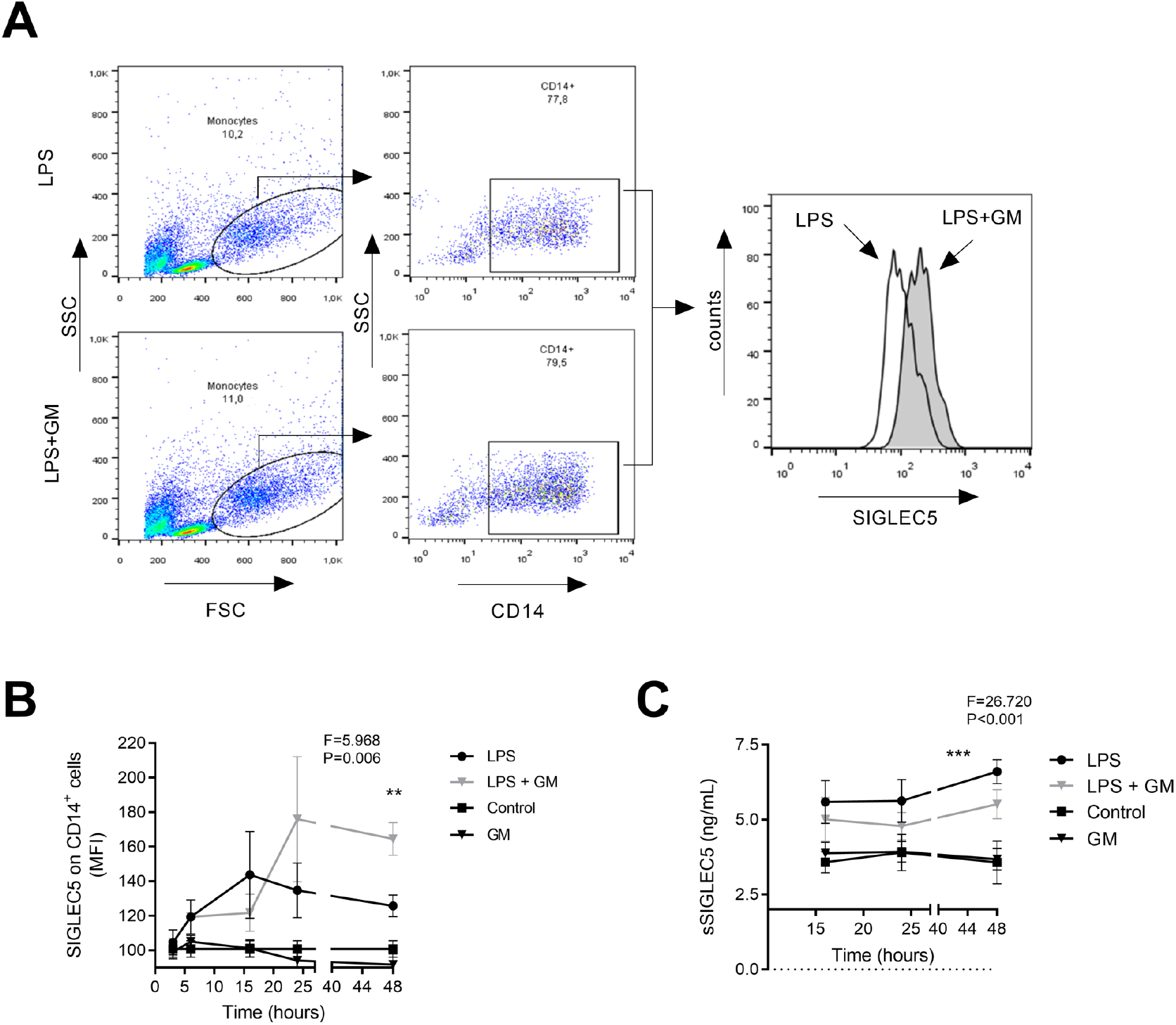
Metalloproteinases shed SIGLEC5 ectodomain from LPS-stimulated human monocytes. **(A)** Proliferation levels (CFSE^dim^) of PWD-stimulated CD4^+^ (sphere) and CD8^+^ (square) cells from HVs in presence of human recombinant sSIGLEC5r for 5 days (n = 5) (left panel). Representative histograms overlays of the proliferating CFSE^dim^ PWD-stimulated CD4^+^ (central panel) and CD8^+^ (right panel) cells from HVs in presence (grey) or not (empty) of human sSIGLEC5r for 5 days (see gating strategy in **Fig. S4A**). **(B)** Fold change of apoptotic (PI^+^/AnnexinV^+^) CD8^+^ cells isolated from HV and incubated or not with sSIGLEC5r for 24 hours (n = 5). **(C)** Proliferation levels (CFSE^dim^) of CD8^+^ cells from HVs, stimulated or not with PWD and co-cultured for 5 days with autologous monocytes pre-challenged (grey columns) or not (empty columns) with 10 ng/mL of LPS for 16 hours. In some indicated conditions, a blocking antibody against SIGLEC5 (α-SIGLEC5) or an unspecific IgG was added (n = 3, left panel). Representative histogram overlay of proliferation levels (CFSE^dim^) of PWD-stimulated CD8^+^ cells from HVs, and co-cultured for 5 days with autologous monocytes pre-challenged with 10 ng/mL of LPS for 16 hours, in presence or not of a blocking antibody against SIGLEC5, α-SIGLEC5 (right panel). **(D)** MFI of SIGLEC5 on CD14^+^ cells 16 hours after nucleofection with either siRSIGLEC5 or siRControl (n = 4). **(E)** Proliferation levels (CFSE^dim^) of CD8^+^ cells from HV, stimulated or not with PWD and co-cultured for 5 days with autologous monocytes pre-nucleofected or not with a specific siRNA against SIGLEC5 (siRSIGLEC5) and pre-challenged (grey columns) or not (empty columns) for 16 hours with 10 ng/mL of LPS (n = 4) (left panel). Representative histogram overlay of proliferation levels (CFSE^dim^) of PWD-stimulated CD8^+^ cells from HVs, and co-cultured for 5 days with autologous monocytes pre-challenged with 10 ng/mL of LPS for 16 hours and pre-nucleofected or not with siRSIGLEC5 (right panel). Statistical analysis was performed using paired t-test (**A-E**). \**P*<0.05; \*\**P*<0.01; \*\*\**P*<0.001.

**Supplemental Figure 2.**
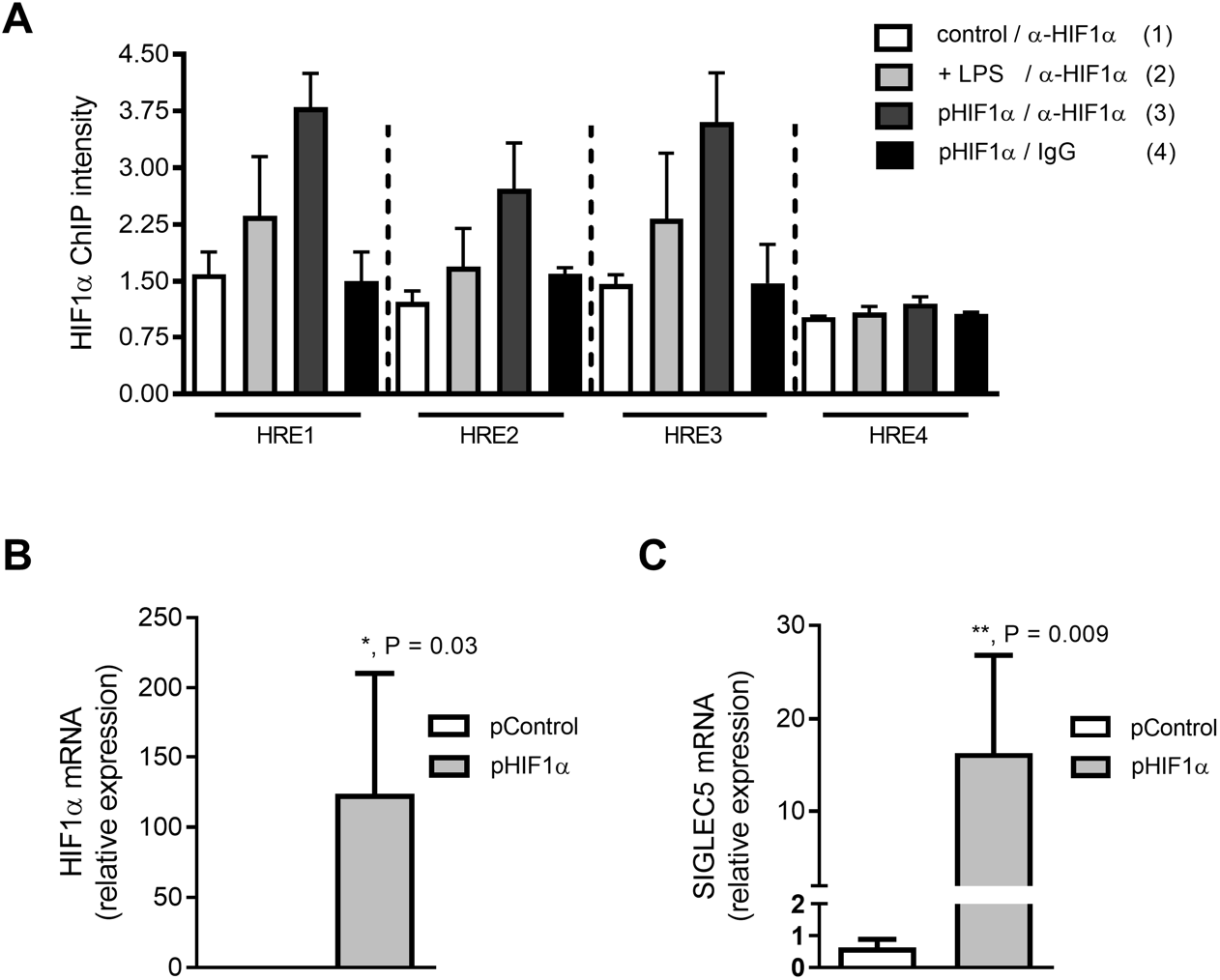
Metalloproteinases shed SIGLEC5 ectodomain from LPS-stimulated human monocytes. **(A)** HREs band intensities of PCR products in the human *SIGLEC5* sequence (Ensembl, ENSG00000105501) based on the consensus sequence [5’-ACGTG-3’] (see **Table S3**) normalised to one of the negative controls (n = 4). **(B)** Relative expression (mRNA) by RT-qPCR of HIF1α on CD14^+^ cells from HV nucleofected with pHIF1α or a control plasmid (n = 6). **(C)** Relative expression (mRNA) by RT-qPCR of SIGLEC5 mRNA HV nucleofected with pHIF1α or a control plasmid (n = 6). Statistical analysis was performed using paired t-test (**B** and **C**). \**P*<0.05; \*\**P*<0.01.

**Supplemental Figure 3.**
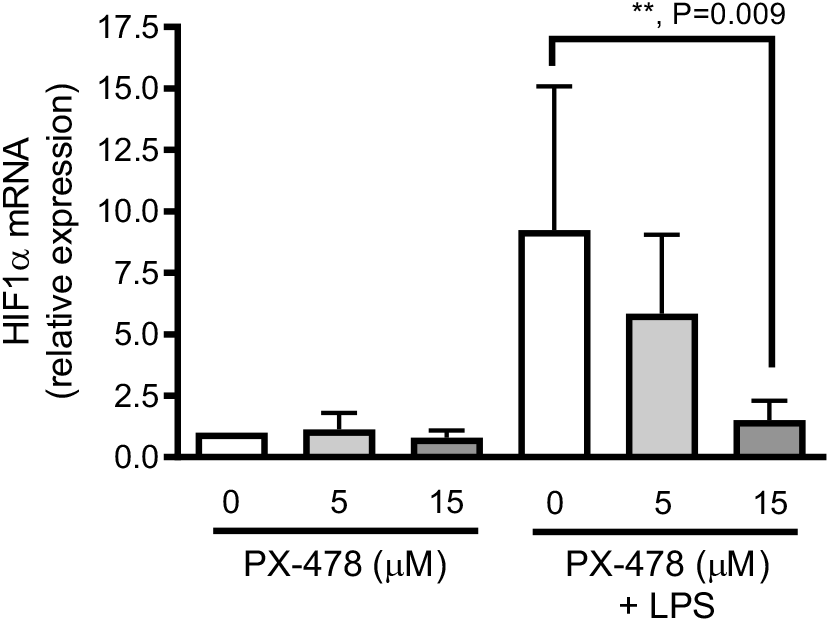
PX-478 inhibits HIF1α expression in human monocytes. Relative expression (mRNA) by RT-qPCR, of HIF1α on CD14^+^ cells from HV, pre-treated or not with different concentrations of a specific inhibitor of HIF1α (PX-478) for 3 hours, and then challenged or not with LPS (10 ng/mL) for 16 hours is shown (n = 7). Statistical analysis was performed using paired t-test. \*\**P*<0.01.

**Supplemental Figure 4.**
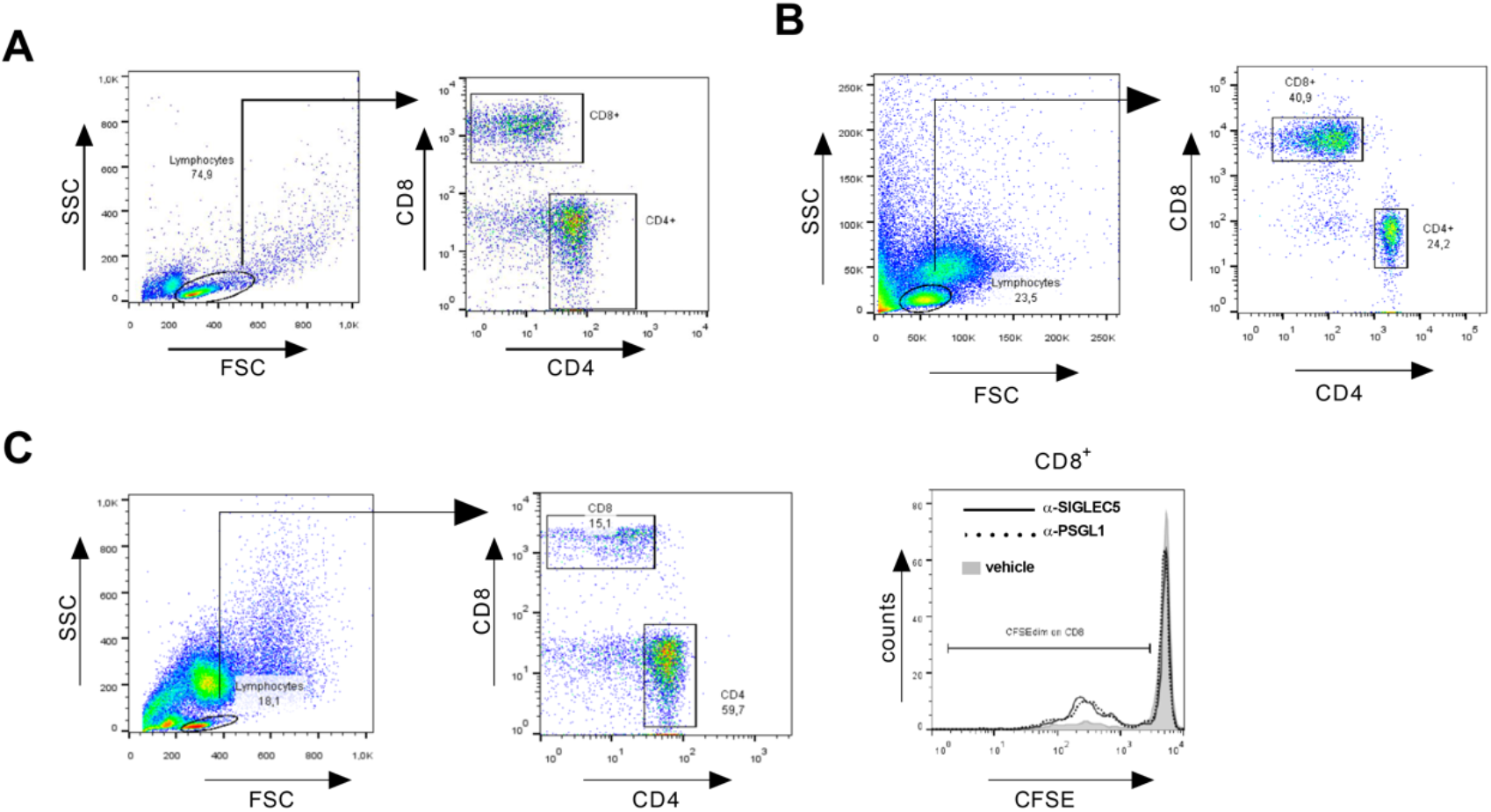
Gating strategy and representative overlay. **(A)** Flow cytometry gating strategy for both CD4^+^and CD8^+^ cells from HV. **(B)** Gating strategy for sSIGLEC5r-FC binding on CD8^+^cells isolated from patients with sepsis. **(C)** Gating strategy for T-cells proliferation from patients with sepsis (left and central panels). Representative histogram overlay of CD8^+^ cells proliferation in presence or not of anti-SIGLEC5 (black line) and anti-PSGL1 (dashed line) blocking antibodies (right panel).

**Supplemental Figure 5.**
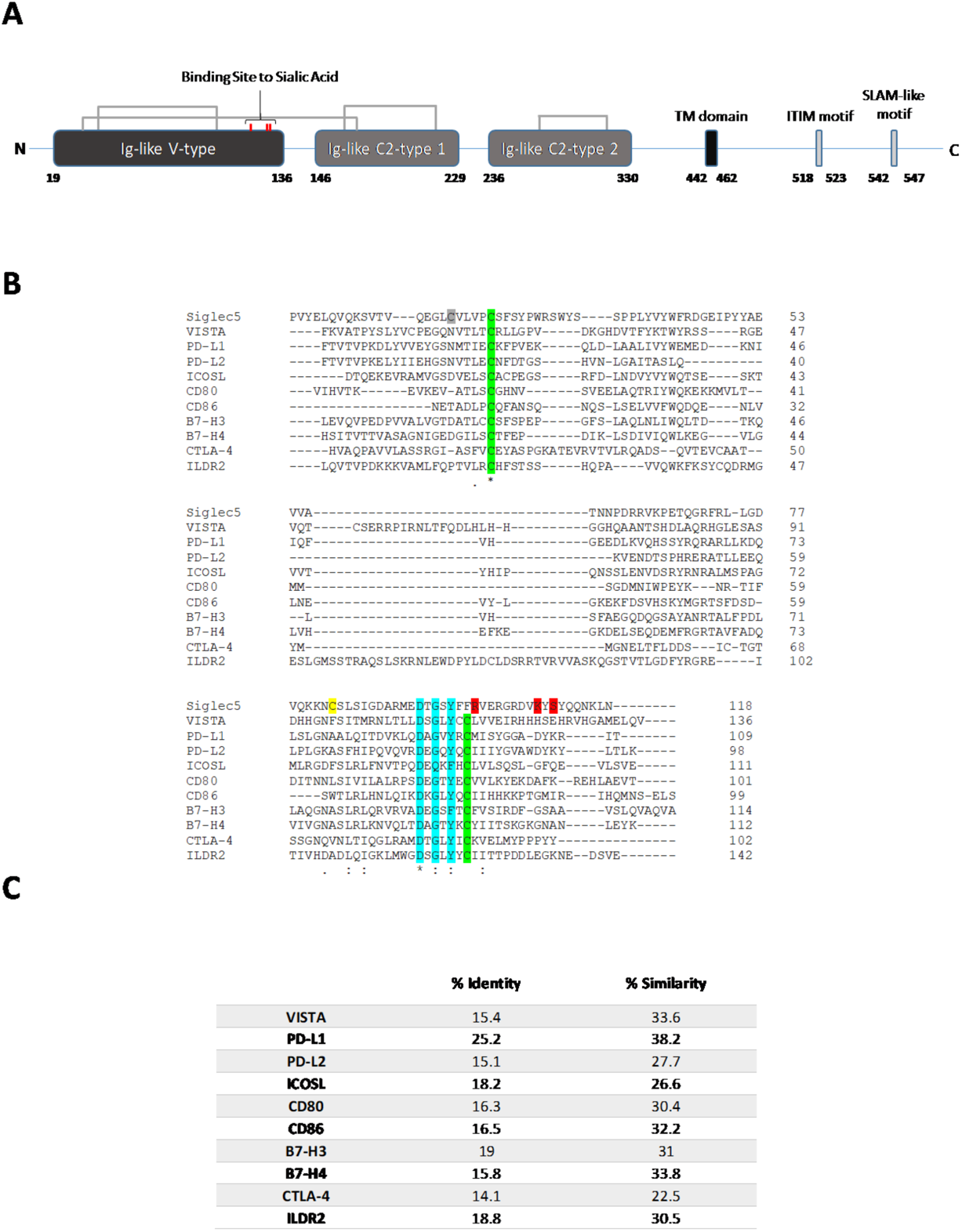
Human SIGLEC5 exhibits Ig-like-V-type regions and high similarity to other immune checkpoints. **(A)** Schematic representation of human SIGLEC5 protein primary structure. Black and grey boxes represent the main domains, interconnecting lines represent disulphide bridges, red ticks on the Ig-like-V-type domain represent the amino acids responsible for the sialic acid binding. The transmembrane domain (TM) is also displayed. **(B)** Multiple sequence alignment of the Iglike-V-type domains of SIGLEC5 and other proteins postulated and confirmed as immune checkpoints. DxGxYxC motifs are highlighted in blue and the amino acids responsible for the sialic acid binding are highlighted in red. Canonical and non-canonical cysteine implicated on the inter-disulphide bridge are represented in green and yellow, respectively. **(C)** Percentages of identity and similarity between the Ig-like-V-type domains of SIGLEC5 and other proteins postulated and/or confirmed as immune checkpoints.

**Supplemental Figure 6.**
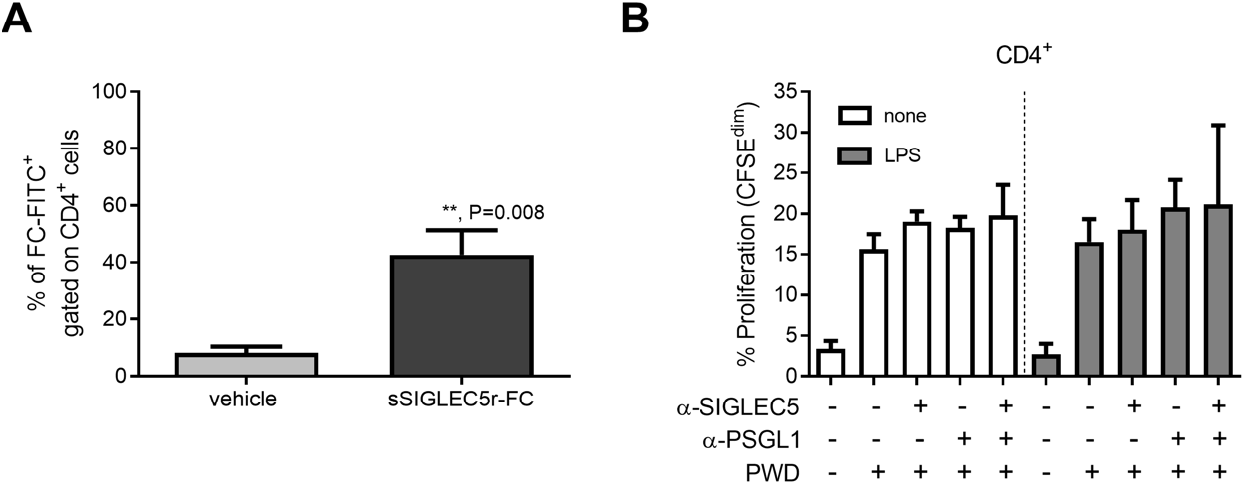
SIGLEC5 binds to CD4^+^ cells from patients with sepsis but does not impair their proliferation. **(A)** Quantification of the binding of sSIGLEC5r-FC to CD4^+^ cells from septic patients (n = 4). The binding was revealed by an antibody (α-FC-FITC). **(B)** Proliferation levels (CFSE^dim^) of CD4^+^ cells from septic patients in presence or not of PWD and co-cultured for 5 days with autologous monocytes pre-challenged (grey columns) or not (empty columns) with 10 ng/mL of LPS for 16 hours. In some conditions, the α-PSGL1 and α-SIGLEC5 antibodies were added (n = 6). Statistical analysis was performed using paired t-test (**A** and **B**). *\*\*P*<0.01.

**Supplemental Figure 7.**
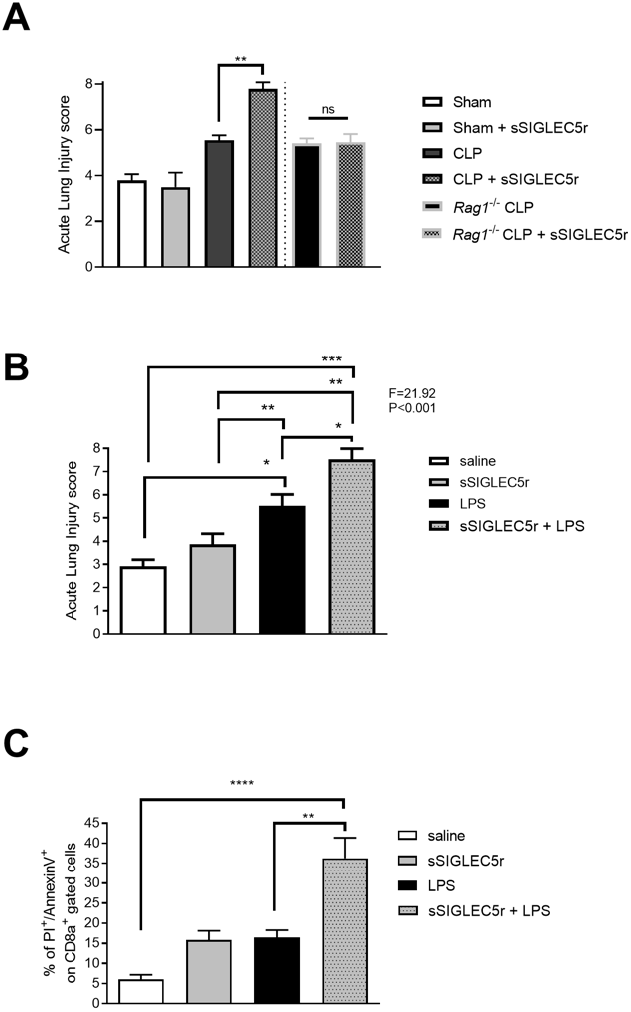
SIGLEC5 induces lung injury in animal models of CLP and endotoxemia. **(A)** Acute lung injury evaluated on Haematoxylin/Eosin stained lung sections of mice from indicated groups. **(B)** Acute lung injury evaluated on Haematoxylin/Eosin stained lung sections of mice from indicated groups. **(C)** Quantification of late apoptosis of mice CD8a^+^-splenocytes measured by flow cytometry. Statistical analysis was performed using unpaired t-test (**A** and **C**) or a one-way ANOVA with Tukey’s multiple comparisons test. *, *P*<0.05; **, *P*<0.01; ***, *P*<0.001; ****, *P*<0.0001.

**Supplemental Table 1.** Description of the patients with Aneurysm or Stroke, and Healthy Volunteers.

| Characteristic* | Aneurysm<br>(N=11) | Stroke<br>(N=16) | Healthy Volunteers<br>(N=100) |
| --- | --- | --- | --- |
| Age (years) | 62±6.88 | 70±13.61 | 50±11.98 |
| Male sex – n (%) | 5(38.46) | 10(58.82) | 54(54) |
| APACHE II | 11.08±7.68 | - | - |
| CRP, mg/L | 18.19±30.70 | 9.56±11.36 | - |
| INR | 1.02±0.06 | 1.08±0.26 | - |

\* Data are presented as mean±SD
APACHE II: Acute Physiology and Chronic Health Evaluation II; CRP: C-reactive protein; INR: international normalised ratio.

**Supplemental Table 2.**
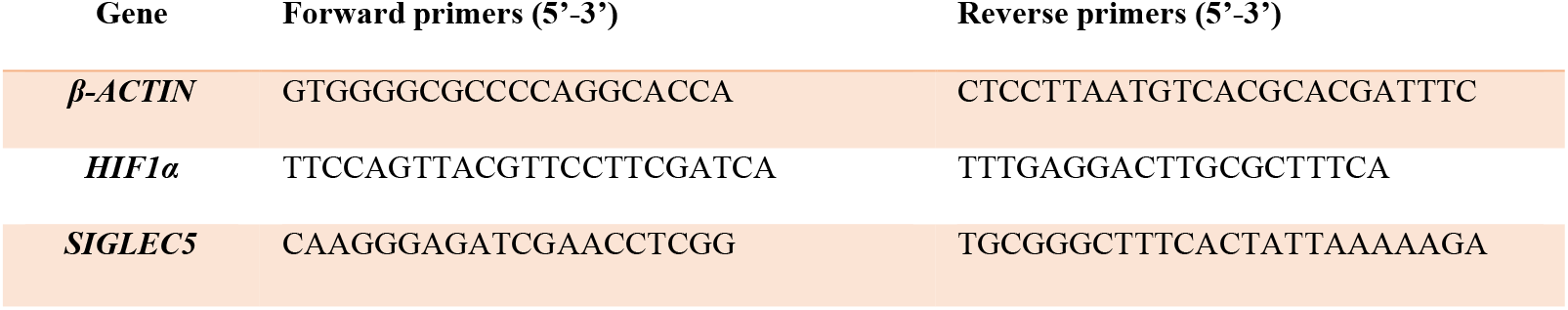
Sequence of primers used in RT-pPCR2.

**Supplemental Table 3.**
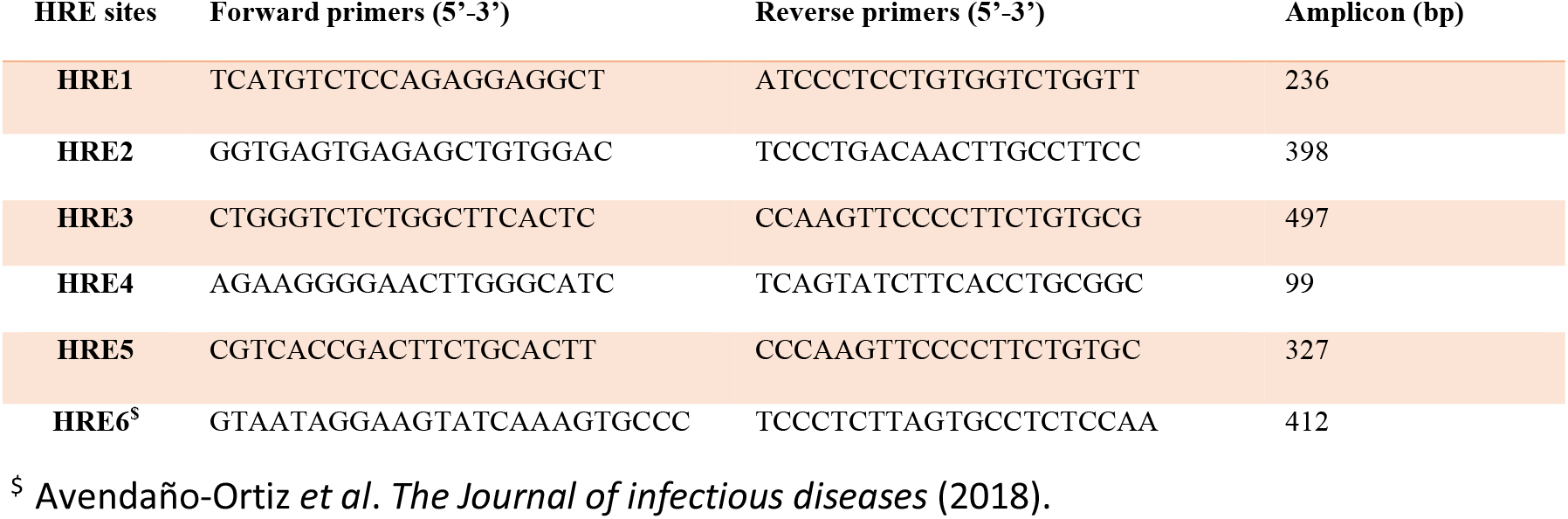
Sequence of primers used in RT-pPCR2.

